# Neurofilament light protein as a blood biomarker for Huntington’s disease in children

**DOI:** 10.1101/2021.02.02.21251000

**Authors:** Lauren M Byrne, Jordan L Schultz, Filipe B Rodrigues, Ellen van der Plas, Douglas Langbehn, Peg Nopoulos, Edward J Wild

## Abstract

Juvenile-onset Huntington’s disease (JoHD) is a rare, particularly devastating form of Huntington’s Disease (HD) for which clinical diagnosis is challenging and robust outcome measures are lacking. Neurofilament light protein (NfL) in plasma has emerged as a prognostic biomarker for adult-onset HD. We report that plasma NfL is elevated in JoHD and premanifest HD mutation-carrying children. Quantifying plasma NfL may improve clinical diagnosis and therapeutic trial design in the pediatric population.

## Main text

Juvenile-onset HD (JoHD) is a devastating form of Huntington’s Disease (HD) that manifests before the age of 21^1^. While the underlying pathology of CAG repeat expansions in *HTT* exon 1 is similar to adult-onset HD (AoHD), JoHD has typically much larger expansions. JoHD has unique disease features including prominent rigidity, dystonia, and bradykinesia, and tends to progress rapidly. There are no disease-modifying therapies for HD but numerous targeted therapeutics are in advanced trials^2^. JoHD is ultra-rare (approximately 5 per million^3^), contributing to a lack of robust clinical scales or biomarkers that could facilitate clinical trials in this population. Further, initial manifestations of JoHD often overlap with normal variability in childhood or adolescence, or with prevalent juvenile disorders such as depression, anxiety, ADHD, Tourette’s syndrome, complicating difficult decisions about diagnosis and genetic testing of minors^1^.

Similar to other neurological diseases, plasma neurofilament light chain protein (NfL) has emerged as a biomarker for AoHD that strongly correlates with clinical, cognitive and MRI measures^4^. Baseline plasma NfL concentration increases early and predicts subsequent HD progression and brain atrophy^4–7^. In a cohort of young adult premanifest HD (preHD) mutation carriers, on average 24 years from estimated onset, NfL concentrations in CSF and plasma were the earliest detectable alterations in HD^8^.

Despite being a robust biomarker for AoHD^4–7^, NfL has yet to be evaluated in JoHD or in children and adolescent HD mutation carriers who will develop HD in adulthood but have no disease manifestations (preHD). We quantified plasma NfL in a unique cohort of 9 JoHD subjects, 30 preHD and 61 mutation-free control children and adolescents.

Characteristics of the age-matched cohort are in Supplementary Table 1. We assessed potential confounding demographics (age: Extended Data Fig. 1a; parental socioeconomic status: Extended Data Fig. 1b; sex: Extended Data Fig. 1c; Tanner stage [puberty classification system]: Extended Data Fig. 1d), finding little evidence for the impact of healthy development on plasma NfL concentrations.

We investigated whether plasma NfL was elevated in JoHD. JoHD patients had significantly higher plasma NfL than controls (32.46 pg/mL, [95% Confidence interval (95%CI) 18.08 – 58.27], versus 4.76 pg/mL, [95%CI 4.33 – 5.23] respectively; p<0.0001; Fig. 1a). In JoHD, plasma NfL rose with age on a trajectory significantly distinct from controls (Age*Group: 1.09 pg/mL/year [Confidence interval (95%CI) 1.02-1.16], p=0.013; Fig. 1b). A cutoff of 11.1 pg/mg identified 91% of JoHD cases and 96% of controls (AUC 0.96; Fig. 1c). We also evaluated the relationship between plasma NfL and JoHD clinical measures (Supplementary Table 2). There were no associations between plasma NfL and UHDRS total motor scale, the JoHD Motor scale, or its components, with the exception of the Bradykinesia Drinking measure which had a positive association with plasma NfL (LMER Est. 1.25, [95%CI 0.46 – 2.05] p=0.017). However, this did not survive multiple comparisons correction (false discovery rate p=0.103). There was a significant, positive association between disease duration and plasma NfL concentration (LMER Est. 2.18 [95%CI 0.31 – 4.05] p=0.045). No significant associations were found between plasma NfL and disease burden score (DBS) or brain volumes in JoHD.

**Fig. 1.**
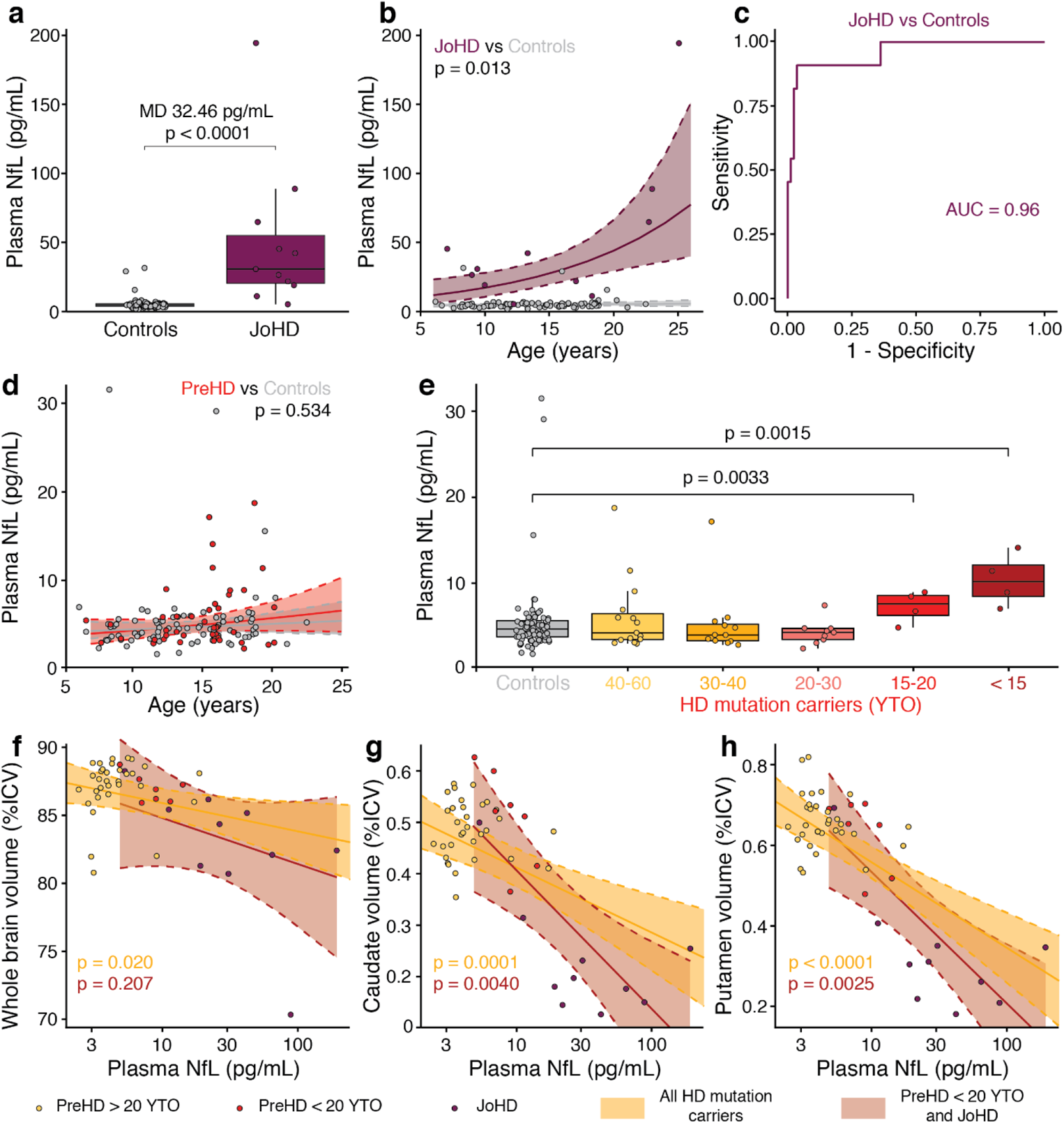
Plasma NfL in JoHD and children at-risk for HD. **a**, Plasma NfL groupwise differences and **b**, modelled plasma NfL trajectories with age in JoHD compared to controls. **c**, Receiver operating characteristic curve for Plasma NfL discriminating JoHD from controls. **d**, modelled plasma NfL trajectories with age in preHD and **e**, YTO groupwise plasma NfL differences compared to control children. Plasma NfL associations with **f**, whole brain **g**, caudate and **h**, putamen volumes in all HD mutation carriers or those with JoHD and within 20 YTO. Data points are raw plasma NfL. Statistics were generated from LMER models with effects per participant and per family (intercepts) and using Satterthwaite approximation for degrees of freedom. P values in e are adjusted for multiple comparisons. For **f-h**, all brain volumes were %ICV; models additionally included a covariate to account for different scanners; and model statistics are presented in Supplementary Table 4. NfL, neurofilament light protein; JoHD, Juvenile-onset Huntington’s disease; preHD, premanifest HD; YTO, years to onset; ICV, intracranial volume; LMER, linear mixed effects regression; MD, mean difference.

Next, we investigated plasma NfL in preHD children. Overall, the age trajectories of NfL in preHD compared to healthy controls were not statistically different from one another (Age*group: 1.02 pg/mL/year [95%CI 0.97-1.06], p=0.534; Fig. 1d). We subdivided preHD by their predicted years to onset (YTO) to assess whether plasma NfL was elevated in children closer to their predicted onset. The characteristics of the YTO groupings are presented in Supplementary Table 3. The mean plasma NfL concentration in controls was 4.76 pg/mL, (95%CI 4.33 – 5.23), which did not differ significantly from preHD 40-60 YTO (5.21 pg/mL, [95%CI 3.96 – 6.84]), 30-40 YTO (4.53 pg/mL, [95%CI 3.45 – 5.94]), or 20-30 YTO (4.10 pg/mL, [95%CI 3.32 – 5.05]). However, the preHD groups that were 15-20 years from onset (7.03 pg/mL [95%CI 5.34 – 9.25]) and within 15 years from predicted onset (10.07 pg/mL [95%CI 7.51 – 13.52]) both had significantly higher plasma NfL than controls (t=3.31, p_FDR_=0.0033 and t=3.72, p_FDR_=0.0015, respectively; Fig. 1e).

In a combined group of HD mutation carriers (preHD+JOHD), we explored the relationship between plasma NfL and brain volume, including whole brain, caudate and putamen volume, which are of primary pathological importance in HD. There were significant negative associations (Supplementary Table 4; Fig. 1f-h) between plasma NfL and whole-brain (LMER Est. −0.90 [CI −1.64 – −0.16] p=0.020; Fig. 1f), caudate (LMER Est. −0.06 [95%CI −0.08 – −0.03] p=0.0001; Fig. 1g) and putamen volumes (LMER Est. −0.09 [95%CI −0.12 – −0.06] p<0.0001; Fig. 1h). When exploring at the associations limiting the analysis to JoHD and preHD children within 20 years from predicted motor onset, the respective slopes of these associations were greater but, given the reduced sample size, only caudate (LMER Est. −0.12 [95%CI −0.18 – −0.05] p=0.0040; Fig. 1g) and putamen (LMER Est. −0.14 [95%CI −0.21 – −0.07] p=0.0025; Fig. 1h) volumes remained significant (Supplementary Table 4).

We combined the KidsHD and KidsJoHD cohorts with HD-CSF – a longitudinal cohort of 80 adult HD mutation carriers and controls – to compare the course of plasma NfL throughout life in health, JoHD and adult-onset Huntington’s disease (Fig. 2a). Plasma NfL initially begins to rise from approximately DBS 200.

**Fig. 2.**
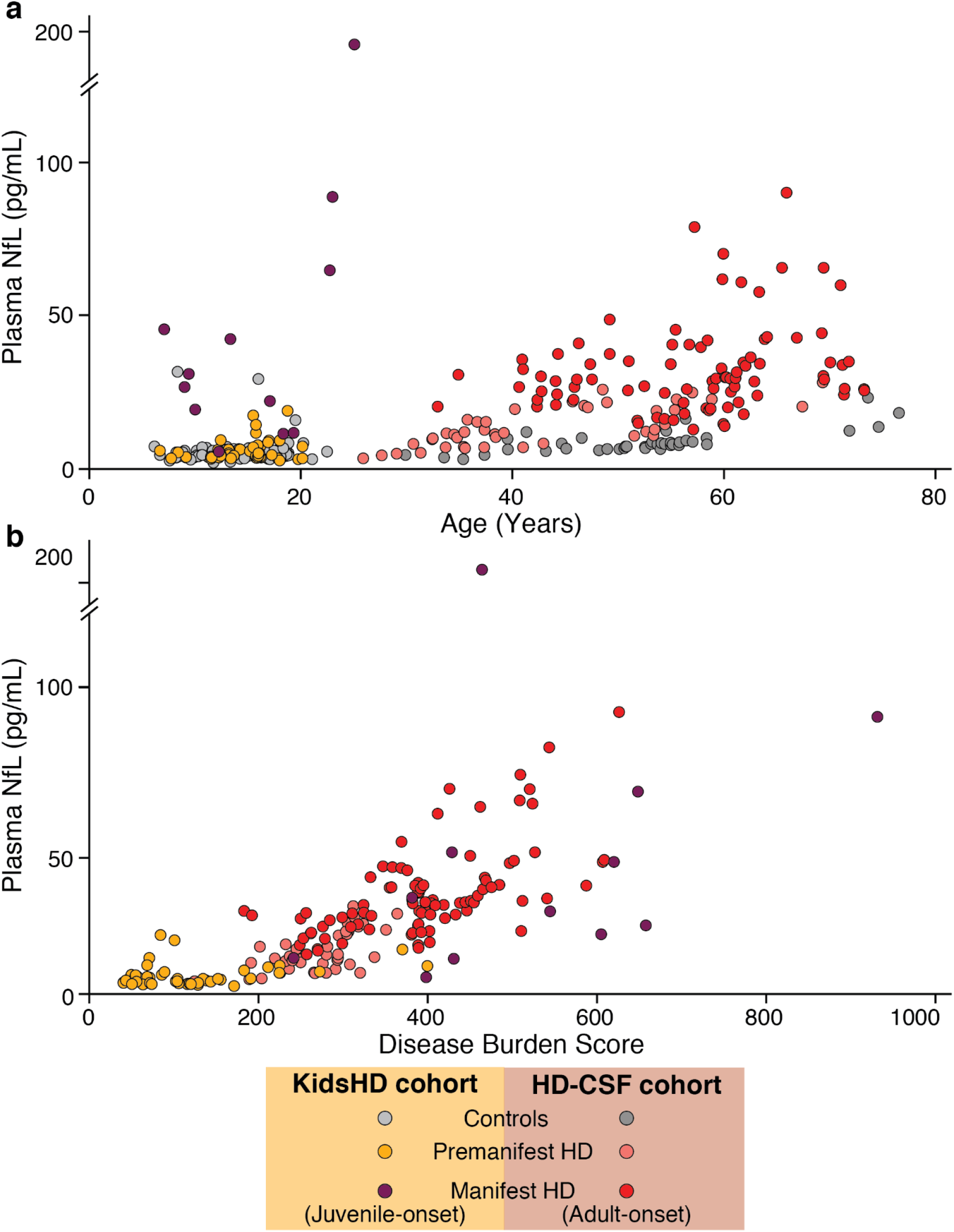
Plasma NfL throughout the HD mutation carrier lifespan. Plasma NfL plotted against **a**. age and **b**. disease burden score in both the Kids cohort and HD-CSF cohort. Data points are raw plasma NfL. NfL, neurofilament light protein.

The elevation and distinct age trajectory of plasma NfL in JoHD patients compared to healthy control children are consistent with those observed in adult onset HD^4,6,7^. Associations with clinical measures were modest, although this may be driven by lack of power and limitations in JoHD clinical measures. Furthermore, as previously reported in AoHD^4–7^, plasma NfL was inversely associated with brain volumes in children with HD mutations, whether JoHD or preHD. Plasma NfL elevation in preHD children within 20 years from predicted onset is consistent with increases reported in a young adult HD cohort. Morphological neurodevelopment differences in the striatum^9^ may limit the utility of MRI to distinguish between the neurodevelopmental and neurodegeneration effects of the HD mutation. NfL could provide insights into the point where neurodevelopment is the predominant concern, potentially helping to inform decisions around the timing of disease-modifying interventions Collectively, these findings suggest two potential applications of NfL: 1) identification of high-risk children for early diagnosis and intervention; and 2) assessing neuroprotection in cohorts of all ages. By evaluating NfL across a wide age range, we can begin to estimate plasma NfL thresholds indicative of neurodegeneration. This could be of value in informing ethically-challenging decisions about which at-risk children showing possible disease manifestations should undergo genetic testing. We conclude that plasma NfL shows promise as a blood biomarker of JoHD and may facilitate clinical trial design in this rare and understudied population.

## Data Availability

The data that support the findings of this study are available from the corresponding author upon reasonable request.

## Extended Data

**Extended Data Fig. 1.**
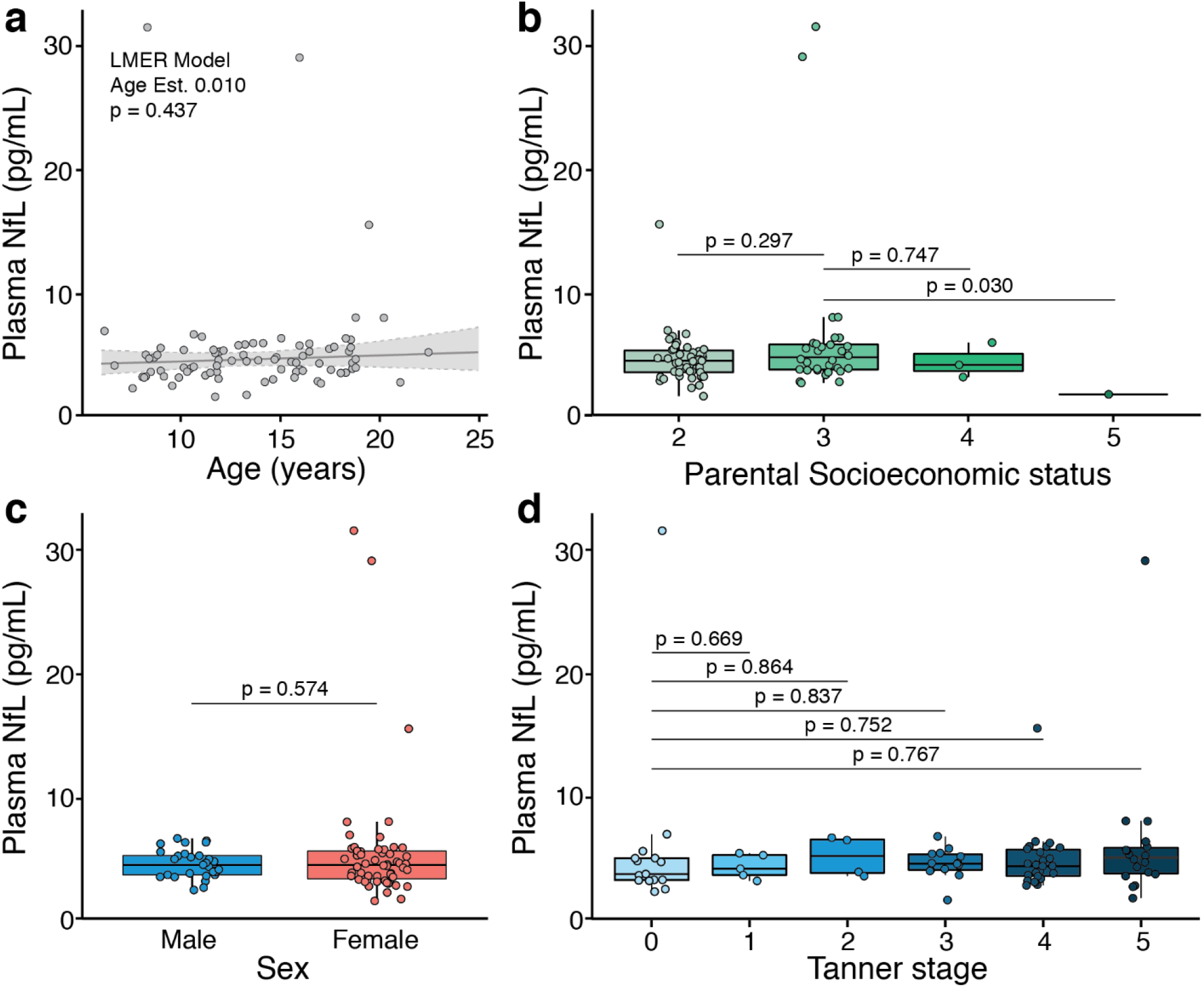
Plasma NfL in healthy development and potential confounding demographics. Plasma NfL concentrations are plotted against **a**, age **b**, parental socioeconomic status **c**, sex and **d**, Tanner stage (puberty) in healthy control individuals (n=61). Data was available from multiple timepoints for some individuals (83 observations). P-values were generated from linear mixed effects regression models with a random participant effect (intercept). Pairwise tests were post-hoc t-tests using Satterthwaite’s method. Raw NfL values are presented. Models used natural log transformed NfL data and was back-transformed for **a**, the age model. NfL, neurofilament light protein; LMER, linear mixed effects regression.

## Acknowledgements

We would like to thank all the participants who took part, providing their time, data and samples to make this work possible.

## Author Contributions

EJW, PN, LMB, JLS and DL were responsible the conception and design of the work; LMB, JLS, FBR and EVP were involved in the acquisition of data; LMB and JLS performed the data analysis; EJW, PN, LMB, JLS and DL were involved in the interpretation of data; LMB drafted the first manuscript draft with JLS and incorporated coauthor revisions. All authors approved the submitted version of the manuscript

## Online methods

### Study design

This retrospective analysis involved quantification of plasma NfL using available blood samples and matching phenotypic data that were collected as part of the KidsHD and KidsJoHD studies – observational studies at the University of Iowa that recruited children and young adults with a family history of HD or with Juvenile Onset HD respectively; as well as healthy control volunteers with no known family history of HD. Recruitment began in May 2009 and was completed in January 2018. All KidsHD/JoHD participants provided either saliva or blood for genetic testing which was for research purposes only; results were not revealed to participants, their families, clinicians, or researchers. Both studies implemented an accelerated longitudinal design where some participants attended multiple visits at 1-2 year intervals and so had repeated measures as previously described^9,10^. The Institutional Review Board at the University of Iowa approved the KidsHD and KidsJoHD studies. The parents or guardians of participants who were less than 18 years old provided written consent and the children provided written assent. Participants who were 18 years or older provided written consent. We combined plasma NfL data from both KidsHD and KidsJoHD with previously published plasma NfL data from the longitudinal HD-CSF cohort to visually assess how NfL fluctuates over the lifespan of the HD mutation carrier. HD-CSF has been extensively described^6,11,12^ (online protocol: DOI: 10.5522/04/11828448.v1).

### Participants

For the current analysis, at-risk participants from Kids-HD were classified as premanifest HD (preHD) mutation carriers if their CAG repeat length was ≥ 36 and healthy controls if <36. JoHD was defined as a UHDRS Total Motor Score ≥ 10 before the age of 21 in the context of the HD mutation and a confirmed diagnosis by a trained neurologist. Plasma samples were available for 30 preHD (44 visits), 60 healthy controls (82 visits) and nine JoHD participants (11 total visits). Clinical severity in JoHD was assessed using the UHDRS total motor score, the JoHD Motor scale^13^ – an adapted version of UHDRS total motor score which weighs JoHD-specific components higher – and disease duration (determined by length of time from clinical diagnosis to time of study visit in years). Disease burden score (DBS) – an arithmetic composite of CAG and age that encompasses the cumulative burden of the CAG expansion [Age * (cAG-35.5)] – was used as a known predictor of HD progression^14^.

The preHD group was divided into subgroups based on their predicted years to motor onset (YTO) into the following groups: 40-60 YTO; 30-40 YTO; 20-30 YTO; 15-20 YTO; <15 YTO. This was calculated using the using their CAG repeat length and age in the formula from Langbehn and colleagues^15^.

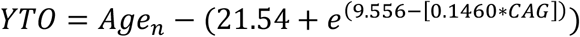

This formula was used only for participants with a CAG repeat length of 36-49. For participants with a CAG repeat length of 50 or above, the Langbehn formula was slightly modified to provide a more accurate estimate of the predicted years to onset. Participants were excluded from either study if they had a history of a major neurologic illness, brain surgery, or significant head trauma.

### Plasma Sample Collection and Storage

KidsHD/JoHD blood samples were collected in EDTA Vacutainer tubes (BD). All samples were coded with a unique study ID and were sent to the University of Iowa Diagnostic Laboratories. Samples were briefly stored on ice and then centrifuged at 2,000g for x minutes. Plasma was decanted before being centrifuged at 3,000g at 4°C for 15 minutes then pipetted into 1.2-mL cryotubes in 500 uL aliquots and frozen and stored at −80°C before being shipped.

### NfL Quantification

KidsHD/JoHD plasma NfL concentrations were measured using the commercially available NF-Light^®^ assay kit from on the Simoa HD-1 analyzer platform as per the manufacturer’s instructions (Quanterix, Lexington). NfL was quantified in duplicate for all plasma samples. The intra-assay coefficient of variation (CV) was 6.5% and inter-assay CV was 1.65%. HD-CSF plasma NfL was previously quantified and published as per Rodrigues et al^7^.

### Image Acquisition and processing

The structural imaging acquisition methods have been described previously^9^. A 3T Siemens Trio TIM (Siemens AG, Munich, Germany) machine was used to acquire images prior to June 2016 (n=33). After June 2016, a 3T General Electric Discovery MR750w (GE Medical Systems, Chicago, IL) was used (n=17). Full details regarding image processing have been previously described by van der Plas, et al^9^.

### Statistical Analysis

In accordance with previous evaluations of plasma NfL, all available NfL concentrations underwent natural log-transformation to account for non-normal distribution^4,6^. We assessed potentially counfounding demographics and measures of healthy development on plasma NfL. None of the variables assessed had an effect on plasma NfL. However, since NfL has been shown to be associated with healhty aging, we included age as a covariate for subsequent analyses^16^. For the primary analyses-of-interest, we performed linear mixed effects regression (LMER) models. The accelerated longitudinal design of KidsHD/JoHD allows for modeling trajectories of development over time while statistically accounting for the correlation that occurs from repeat visits from individuals^9^. All LMER models also included random effects per family to account for siblings in the dataset. P-values were estimated using Satterthwaite approximation for degrees of freedom throughout.

For groupwise comparisons, we compared LMER model adjusted mean plasma NfL values between groups. In these models, within-subject and residual variances were estimated separately for the various groups via iteratively re-weighted least squares. This was done to account for sizable differences in variation between the groups. We adjusted for multiple comparisons using false-discovery-rate (FDR) corrections.

The age*group interaction was evaluated to determine group differences in age-trajectories of plasma NfL. To assess associations with clinical measures, LMER models were constructed with plasma NfL as the independent variable and clinical measures as the dependent variables. LMER models for DBS – a predictor of HD progression – included NfL as the dependent variable. Age was not included as a covariate in either of the models that investigated the relationship between disease duration or DBS and plasma NfL to avoid collinearity, since age was used to calculate these variables. For neuroimaging measures, brain volumes were the dependent variables and NfL independent within the LMER models to permit the addition of scanner as a covariate to account for scanner differences. All brain volumes were presented as percent of intracranial volume (ICV).

We created a receiver operating characteristic curve by performing a generalized binomial regression model with plasma NfL as the independent predictor variable for whether or not a participant was a healthy control or had JoHD. We calculated Youden’s Index by adding the sensitivity and specificity at all available plasma NfL levels. The maximal specificity cutoff point was at 0.964 and the maximal sensitivity cutoff point was at 0.909 and this point correlated with a plasma NfL cutoff point of 11.1 pg/mL. These analyses were performed using the pROC package in RStudio (version 1.16.2)

Groupwise analyses were performed using the PROC MIXED function within SAS (v9.4, SAS Institute Inc, Cary, NC, USA). All other models were performed in RStudio version 1.3.159 using the lmerTest version 3.1.

## Supplementary material

**Supplementary Table 1.** Baseline Cohort Characteristics of KidsHD and KidsJoHD. P-values for continuous variables were generated from one-way analyses of variance. P-values for categorical variables were generated from Pearson Chi-Square analyses. Values are presented as mean ± SD unless otherwise stated. preHD, premanifest Huntington’s disease; JoHD, Juvenile-onset Huntington’s disease. SD, Standard deviation; CAG, Cytosine-Adenine-Guanine; BMI, Body mass index; SES, socioeconomic status, measured by the Hollingshead Scale; NfL, neurofilament light protein.

|  | Controls | preHD | JoHD | P-Value |
| --- | --- | --- | --- | --- |
| **N (total visits)** | 61 (83) | 30 (44) | 9 (11) | N/A |
| **Age** | 12.75 ± 3.71 | 14.00 ± 3.12 | 16.48 ± 6.38 | 0.40 |
| **Males, N (%)** | 23 (37.7) | 10 (33.3) | 5 (55.6) | 0.483 |
| **CAG** | 20.33 ± 4.39 | 43.90 ± 4.47 | 72.11 ± 12.67 | <0.0001 |
| **BMI** | 22.14 ± 7.48 | 21.92 ± 5.13 | 21.51 ± 6.22 | 0.963 |
| **Tanner Stage, N (%)** |  |  |  | 0.337 |
| **0** | 15 (22.2) | 4 (13.3) | 2 (22.2) |  |
| **1** | 4 (6.6) | 0 (0) | 0 (0) |  |
| **2** | 4 (6.6) | 1 (3.3) | 0 (0) |  |
| **3** | 12 (19.7) | 5 (16.7) | 0 (0) |  |
| **4** | 16 (26.2) | 15 (50.0) | 4 (44.4) |  |
| **5** | 10 (16.4) | 5 (16.7) | 3 (33.3) |  |
| **Parental SES, N (%)** |  |  |  | 0.062 |
| **1** | 0 (0) | 0 (0) | 0 (0) |  |
| **2** | 36 (59.0) | 14 (46.7) | 2 (22.2) |  |
| **3** | 22 (36.1) | 14 (46.7) | 4 (44.4) |  |
| **4** | 2 (3.3) | 2 (6.7) | 2 (22.2) |  |
| **5** | 1 (1.6) | 0 (0) | 1 (11.1) |  |
| **Plasma NfL (pg/mL)** | 4.71 ± 1.55 | 5.05 ± 1.57 | 35.16 ± 2.95 | <0.0001 |

**Supplementary Table 2.** Clinical measures and their associations with plasma NfL in JoHD. P-values were generated from linear mixed effects regression models with effects per participant and per family (intercepts) and using Satterthwaite approximation for degrees of freedom. Values are presented as the beta estimate of the model relative to the log-transformed plasma NfL concentration. The models that assessed the relationship between disease duration and disease burden score did not include age as a covariate, as both independent variables are calculated using age. All brain volumes were corrected for intracranial volume and models investigating the relationship between plasma NfL and brain volumes included a covariate to account for different scanners. NfL, neurofilament light protein; UHDRS, Unified Huntington’s Disease Rating Scale; TMS, total motor score; JoHD, Juvenile-onset Huntington’s disease.

| Clinical measures | Association with plasma NfL |  |  |  |
| --- | --- | --- | --- | --- |
|  | N (total visits) | Estimate | Confidence interval | P-value |
| UHDRS TMS | 9 (11) | 7.02 | -13.91 – 27.94 | 0.522 |
| JoHD Motor Scale | 9 (11) | 3.87 | -0.56 – 8.30 | 0.119 |
| Chorea | 9 (11) | 0.07 | -0.57 – 0.71 | 0.832 |
| Bradykinesia Handtapping | 9 (11) | 0.05 | -0.96 – 1.05 | 0.934 |
| Bradykinesia Drinking | 9 (10) | 1.25 | 0.46 – 2.05 | 0.017 |
| Tremors | 9 (11) | 0.41 | -0.38 – 1.19 | 0.333 |
| Disease duration | 9 (11) | 1.19 | 0.03 – 2.35 | 0.045 |
| Disease burden score | 9 (11) | 1.00 | -0.01 – 2.01 | 0.347 |
| Whole Brain Volume | 9 (10) | -3.10 | -7.19 – 0.99 | 0.180 |
| Putamen Volume | 9 (10) | -0.10 | -0.20 – 0.01 | 0.112 |
| Caudate Volume | 9 (10) | -0.07 | -0.15 – 0.01 | 0.116 |

**Supplementary Table 3.** Baseline Cohort Characteristics by years to onset groupings. P-values for continuous variables were generated from one-way analyses of variance. P-values for categorical variables were generated from Pearson Chi-Square analyses. Values are presented as mean ± SD unless otherwise stated. SD, Standard deviation; CAG, Cytosine-Adenine-Guanine; YTO, years to onset; BMI, Body mass index; SES, socioeconomic status, measured by the Hollingshead Scale; NfL, neurofilament light protein.

|  | Controls | 40-60 YTO | 30-40 YTO | 20-30 YTO | 15-20 YTO | <15 YTO | P-Value |
| --- | --- | --- | --- | --- | --- | --- | --- |
| **N (total visits)** | 61 (83) | 9 (15) | 10 (12) | 7 (9) | 2 (4) | 2 (4) | NA |
| **Age** | 12.75 ± 3.71 | 13.99 ± 2.07 | 13.47 ± 3.93 | 13.33 ± 2.98 | 15.04 ± 2.65 | 17.96 ± 3.12 | 0.355 |
| **Female, N (%)** | 23 (37.7) | 4 (44.4) | 8 (80.0) | 6 (85.7) | 1 (50.0) | 1 (50.00) | 0.488 |
| **CAG** | 20.33 ± 4.39 | 39.78 ± 0.97 | 42.60 ± 0.84 | 46.71 ± 2.29 | 49.00 ± 1.41 | 54.00 ± 7.07 | <0.0001 |
| **YTO** | N/A | 50.39 ± 6.23 | 36.38 ± 2.03 | 24.392 ± 2.87 | 17.96 ± 0.79 | 11.06 ± 1.93 | <0.0001 |
| **BMI** | 22.14 ± 7.48 | 22.63 ± 5.88 | 20.68 ± 4.83 | 21.99 ± 4.86 | 18.23 ± 1.16 | 28.42 ± 1.30 | 0.295 |
| **Tanner Stage** |  |  |  |  |  |  | 0.280 |
| 0 | 15 (22.2) | 0 (0.0) | 3 (30.0) | 1 (14.3) | 0 (0.0) | 0 (0.0) |  |
| 1 | 4 (6.6) | 0 (0.0) | 0 (0.0) | 0 (0.0) | 0 (0.0) | 0 (0.0) |  |
| 2 | 4 (6.6) | 1 (11.1) | 0 (0.0) | 0 (0.0) | 0 (0.0) | 0 (0.0) |  |
| 3 | 12 (19.7) | 4 (44.4) | 0 (0.0) | 0 (0.0) | 1 (50.0) | 0 (0.0) |  |
| 4 | 16 (26.2) | 4 (44.4) | 5 (50.0) | 4 (57.1) | 1 (50.0) | 1 (50.0) |  |
| 5 | 10 (16.4) | 0 (0.0) | 2 (20.0) | 2 (28.6) | 0 (0.0) | 1 (50.0) |  |
| **Parental SES, N (%)** |  |  |  |  |  |  | 0.366 |
| 0 | 0 (0) |  |  |  |  |  |  |
| 1 | 36 (59.0) | 5 (55.6) | 7 (70.0) | 1 (14.3) | 1 (50.0) | 0 (0.0) |  |
| 2 | 22 (36.1) | 3 (33.3) | 3 (30.0) | 5 (71.4) | 1 (50.0) | 2 (100.0) |  |
| 3 | 2 (3.3) | 1 (11.1) | 0 (0.0) | 1 (14.3) | 0 (0.0) | 0 (0.0) |  |
| 4 | 1 (1.6) |  |  |  |  |  |  |
| **Plasma NfL** | 4.71 ± 1.55 | 4.81 ± 1.55 | 4.81 ± 1.67 | 4.43 ± 1.32 | 6.42 ± 1.48 | 9.97 ± 1.63 | 0.211 |

**Supplementary Table 4.** Plasma NfL associations with brain volume in HD mutation carriers. P-values were generated from linear mixed effects regression models with random effects per participant and per family (intercepts), age as a covariate and using Satterthwaite approximation for degrees of freedom. Values are presented as the beta estimate of the model relative to the log-transformed plasma NfL concentration. All brain volumes were corrected for intracranial volume and models investigating the relationship between plasma NfL and brain volumes included a covariate to account for different scanners. NfL, neurofilament light protein; HD, Huntington’s disease; JoHD, Juvenile-onset Huntington’s disease.

| Clinical measures | Association with plasma NfL - All HD mutation carriers |  |  |  | Association with plasma NfL – JoHD and within 20 YTO |  |  |  |
| --- | --- | --- | --- | --- | --- | --- | --- | --- |
|  | N (total visits) | Estimate | Confidence interval | P-value | N (total visits) | Estimate | Confidence interval | P-value |
| **Whole Brain Volume** | 35 (49) | -0.90 | -1.64 – -0.16 | 0.020 | 12 (17) | -1.46 | -3.67 – 0.74 | 0.207 |
| **Caudate Volume** | 35 (49) | -0.06 | -0.08 – -0.03 | 0.0001 | 12 (17) | -0.12 | -0.18 – -0.05 | 0.0040 |
| **Putamen Volume** | 35 (49) | -0.09 | -0.12 – -0.06 | <0.0001 | 12 (17) | -0.14 | -0.21 – -0.07 | 0.0025 |

